# Diagnostic accuracy in detecting malignancy in suspicious skin lesions using Artificial Intelligence

**DOI:** 10.1101/2025.03.11.25323753

**Authors:** Miguel Sánchez-Viera, Alfonso Medela, Isabel del Campo, Jordi Barrachina, Concetta D’Alessandro, Alejandra Vallejos, Alejandra Capote, Andy Aguilar, Taig Mac Carthy, Gerardo Fernández, Antonio Martorell

**Author notes:** **Corresponding author:** Alfonso Medela, Department of Medical Computer Vision and PROMs, Legit.Health, Calle Gran Vía Diego López de Haro, 1, 48001 Bilbao, Spain. **Funding:** This study was co-funded by the European Union (NextGenerationEU) through the Public Business Entity Red.es (State Secretariat for Digitalization and Artificial Intelligence, Ministry of Economic Affairs and Digital Transformation) within the framework of the 2021 Call for Grants for research and development projects in artificial intelligence and other digital technologies, and their integration into value chains (C005/21-ED), with file number 2021/C005/00154001. **Ethical approval:** Ethics committee of HM Hospitales (Comité Ético de Investigación con medicamentos) gave ethical approval for this work, under reference number 24.12.2266-GHM. **Ethical statement:** The patients in this manuscript have given written informed consent to participate in this study. **Data availability statement:** All data produced in the present study are available upon reasonable request to the authors.

## Abstract

**Background:** Artificial Intelligence (AI) has demonstrated a high image processing capacity and improved diagnostic accuracy in dermatology. In this context, Computer-Aided Diagnosis (CAD) systems have shown a diagnostic performance comparable to that of specialists in classifying skin lesions, particularly pigmented lesions. The present study aims to validate Legit.Health is a reliable tool for diagnosing and assessing the severity of patients with skin lesions suspicious of malignancy.

**Objective:** To validate that the Legit.Health medical device optimises clinical workflow by enhancing diagnostic accuracy and determining the malignancy or severity of patients with skin lesions suspicious of malignancy.

**Methods:** An observational, prospective study was conducted, incorporating both longitudinal and retrospective cases. A total of 76 retrospective patients with 88 lesions and 32 prospective patients with 42 lesions attending Instituto de Dermatologia Integral Madrid, Spain, were recruited. The diagnostic performance of Legit.Health was compared with that of dermatologists in the retrospective images against a gold standard (biopsy results). In the prospective phase of the study, the performance of the current Legit.Health medical device was evaluated alongside dermatologists assisted by the device and the latest version of the device (Legit.Health Plus). Analyses were performed to calculate the AUC (area under the curve), accuracy, sensitivity, and specificity.

**Results:** In the retrospective analysis, the device demonstrated an AUC of 0.76 compared to 0.79 for dermatologists in detecting malignant lesions. For these images, the device achieved the following accuracy scores: top-1 = 0.23, top-3 = 0.38, and top-5 = 0.47, whereas dermatologists achieved top-1 = 0.33 and top-3 = 0.45 (providing only three possible diagnoses). When the specific histologic subtype of naevus was not considered in the diagnosis, Legit.Health achieved an accuracy of top-1 = 0.50, top-3 = 0.71, and top-5 = 0.78, compared to dermatologists’ top-1 = 0.50 and top-3 = 0.70. In the prospective analysis, we examined the performance of dermatologists using the Legit.Health medical device, the device alone, and the latest version of the device. In the malignancy analysis, they achieved an AUC of 0.94, 0.95, and 0.97, respectively. Regarding diagnostic accuracy, dermatologists assisted by the medical device achieved a top-1 accuracy of 0.30, while both the medical device alone and its latest version achieved top-1 accuracies of 0.22 and 0.26, respectively, which increased to 0.44 and 0.52 when expanding to top-5. When the specific histologic subtype of naevus was not considered in the diagnosis, accuracies increased to 0.85, 0.74, and 0.81, respectively, further improving as top-K was increased to top-5, reaching 0.89 and 0.93, respectively.

**Conclusions:** The device’s diagnostic capability in distinguishing malignant conditions is on par with that of expert dermatologists. This confirms its reliability as a tool for detecting skin malignant categories in ICD-11, assisting in prioritising patients based on urgency and directing them to the appropriate specialist or consultation.

## INTRODUCTION

Image-based Artificial Intelligence (AI) has significant potential to enhance the accuracy of visual diagnosis in the medical field^1,2^. During the COVID-19 pandemic, limited access to in-person healthcare services drove changes in medical care, accelerating the adoption of telemedicine^3^. In this context, AI-assisted triage and decision support can be essential in helping healthcare professionals manage workload and improve efficiency^4,5^.

In dermatology, pigmented lesions are among the most common conditions, generating high demand in dermatology centres^6^. The triage, clinical assessment, and follow-up of these patients require in-person resources and the dedicated time of specialists and their teams. The implementation of AI-driven tools could support these professionals in streamlining such processes and optimising workload management.

Advances in image recognition and interpretation, as well as AI, have driven innovations in diagnosing various conditions, including dermatological diseases. Computer-aided diagnosis (CAD) systems and other algorithm-based technologies have demonstrated their ability to classify lesion images with a performance comparable to that of expert clinicians^7,8^.

Thus, progress in image recognition and AI has led to innovations in the diagnosis of skin lesions^2,9^. With appropriate development and rigorous evaluation, this technology could enhance diagnostic accuracy. Studies have shown that AI algorithms can classify images of lesions, including melanoma, with an expertise level comparable to that of dermatologists^10^.

The advent of computer vision has revolutionised the identification of dermatological conditions, representing a major opportunity to improve the diagnostic process^11,12^. Among the key trends that have contributed to improved patient survival rates are preventive measures and early diagnosis campaigns^13^. Therefore, based on our previous research, we propose an AI-based system to assess the malignancy of skin lesions and optimize diagnosis.

This study will evaluate the Legit.Health medical device, an AI-powered tool designed to optimise clinical workflow and patient management for dermatological conditions. The purpose of this system is to automatically prioritise patients with greater urgency, determine the appropriate type of consultation (dermatological or aesthetic), enhance diagnostic capacity, assist auxiliary staff in detecting malignant pigmented lesions, and provide a visual record (photographic evidence) of the condition for subsequent expert review.

The primary objective of this study is to validate whether the Legit.Health medical device improves efficiency in clinical workflow and patient management for individuals with dermatological conditions. This will be achieved by assessing the tool’s capability to diagnose and determine the severity of skin lesions suspicious of malignancy. Ultimately, this will lead to a reduction in the need for in-person consultations and lower patient care costs by ensuring that cases are directed to the appropriate consultation type from the outset.

## MATERIALS AND METHODS

### Study Design

A prospective observational study with both longitudinal and retrospective case series was conducted, beginning on 2 February 2024 and concluding on 23 August 2024. Patients with lesions suspected of skin malignancy were recruited from the IDEI dermatology centre. Demographic data and image-processing data were obtained using the Legit.Health medical device. The study protocol was approved by the Research Ethics Committee for Medicines at HM Hospitals, under reference: 24.12.2266-GHM.

### Study population

All included patients were adults (≥18 years old) with lesions suspected of skin malignancy. Prospective patients were recruited during their dermatology consultation at IDEI. No medications or additional treatments were administered as part of this study. Retrospective images were selected from patients with histologically studied lesions (both benign and malignant, pigmented and non-pigmented). Retrospective data provided insights into historical diagnostic patterns, whereas prospective data reflected more recent cases, allowing assessment of the device’s utility in real-time clinical scenarios.

### Data collection and outcome measures

Patients were recruited during their dermatology consultation at IDEI. Those who met the inclusion criteria were enrolled in the study. A photograph was taken of the lesion prompting the consultation; if a patient had multiple lesions requiring dermatological examination, images were captured for each lesion. For retrospective patients dermoscopic and clinical images were included, using those with the best quality. For prospective patients, the following data were recorded: age, sex, the clinical diagnosis made by the investigator, the recommended course of action (lesion excision, follow-up, or referral for a cosmetic treatment when no suspicious of malignancy), the histopathological diagnosis (if available), malignancy scoring assigned by the investigator (scale of 1 to 10) and by Legit.Health (scale of 1 to 100), as well as the top-1, top-3, and top-5 diagnostic predictions from Legit.Health. It is important to highlight that in the prospective study, only lesions that appeared suspicious to the clinical or to Legit.Health were excused. Therefore, non-suspicious lesions were not excised and we do not have histological confirmation. However, these lesions were included in the concordance analysis between the clinical diagnosis and that of the device..

For retrospective cases, in addition to these data, the top-1 and top-3 diagnoses provided by the investigator (up to three possible diagnoses) and confirmation of malignancy were also recorded. For prospective patients, all images used were clinical, while for retrospective patients dermoscopic and clinical images were collected in the study.

### Photo capture Procedure

All skin lesions were photographed following these technical guidelines:

- Uncompressed image formats such as PNG, HEIC, or TIFF
- Taken with a smartphone equipped with a minimum camera resolution of 13 megapixels
- All images had post-processing features disabled, including HDR, portrait mode, colour filters, and digital zoom.
- Dermoscopic images were captured using Fotofinder medicam 1000, videodermatoscope from FotoFinder Systems GmbH3Gen Inc.

### Statistical analysis

For diagnostic performance evaluation, histopathological examination served as the reference standard in both retrospective and prospective studies. Various statistical analyses were conducted to assess and compare the performance of dermatologists and the medical device.

The area under the curve (AUC) was calculated to first evaluate the diagnostic performance of dermatologists and the medical device in detecting malignancy, and second to compare the performance of dermatologists assisted by the Legit.Health medical device, the Legit.Health device alone, and the latest version of the device. Sensitivity and specificity were also computed.

For skin lesion recognition, top-K accuracy was calculated. This metric measured the frequency with which the correct diagnosis was among the device’s or dermatologists’ top predictions. It was assessed for dermatologists, the Legit.Health device, and the latest version of the medical device (Legit.Health Plus). For this study, K values of 1, 3, and 5 were used. Differences in diagnostic performance between retrospective and prospective studies were evaluated, attributing improvements to the homogeneity of the prospective lesion dataset and the assistance provided by the medical device.

## RESULTS

### Dataset

The dataset included 88 images from 76 different retrospective patients, of which 77 were dermatoscopic and 11 were clinical images, including for the analysis only the best quality images, clinical or dermoscopic.. Each lesion had a single corresponding image.

The dataset also included 120 images from 42 lesions in 32 different prospective patients. Each lesion had up to three images. All prospective images were clinical. Prospective lesions were also captured alongside the dermatologist’s recommendation regarding their excision.

### Retrospective analysis

The malignancy results demonstrate a similar diagnostic performance between dermatologists and the medical device. Specifically, dermatologists and the medical device achieved an AUC of 0.79 and 0.76, respectively. In terms of sensitivity and specificity, dermatologists reached a sensitivity of 86% and a specificity of 36%, while the medical device recorded a sensitivity of 81% and a specificity of 52%. These values were derived using a threshold of 10, as recommended by the manufacturer. The malignancy results indicate a greater tendency among dermatologists to diagnose malignant pathologies. The confirmed diagnostic data from this part of the study are presented in Supplementary Table 1.

**Supplementary Table 1.** Diagnosed diseases in retrospective study patients and their percentage within the 86 analysed samples.

| Condition | Lesions (n, %) |
| --- | --- |
| Seborrheic keratosis | 15 (17.44) |
| Basal cell carcinoma | 14 (16.3) |
| Junctional melanocytic nevus | 12 (14.0) |
| Compound nevus | 9 (10.4) |
| Nevus | 8 (9.3) |
| Intraepidermal carcinoma | 5 (5.8) |
| Junctional nevus | 5 (5.8) |
| Actinic keratosis | 4 (4.6) |
| Léntigo | 4 (4.6) |
| Benign keratosis | 3 (3.5) |
| Blue nevus | 2 (2.3) |
| Melanoma | 2 (2.3) |
| Non-specific lesion | 2 (2.3) |
| Dermatofibroma | 1 (1.2) |

For the pathological diagnosis analysis, two samples from the dataset were excluded due to the absence of a clear pathological result. The analysis revealed comparable performance between dermatologists and the medical device, as shown in Table 1. It is important to note that for this evaluation, dermatologists provided up to three alternative diagnostic results.

**Table 1.** Diagnostic accuracy results from dermatologists and medical device.

|  | Top-1 Accuracy | Top-3 Accuracy | Top-5 Accuracy |
| --- | --- | --- | --- |
| **Dermatologist** | 0.33 | 0.45 | - |
| **Medical device** | 0.23 | 0.38 | 0.47 |

Upon reviewing the set of diagnoses, it was observed that 36 of the 86 images (42%) corresponded to different histologic subtypes of naevi. Among these, dermatologists and medical device misclassified the specific histologic type of naevus (ie.: Junctional nevus, compound nevus, …) in 24 and 27 out of the 36 cases, respectively.

To provide a broader view of diagnostic performance, the evaluation criteria were adjusted to consider whether any diagnosis of a nevus as correct when a nevus was identified, regardless of its specific type. With this generalised approach, the number of misclassifications was reduced to 2 for dermatologists and 0 for the medical device. This adjustment led to a significant improvement in performance for both, with enhanced top-K accuracy and even a top-5 accuracy for the medical device surpassing that of dermatologists. The results are presented in Table 2.

**Table 2.** Diagnostic accuracy results for dermatologists and the medical device after grouping different types of naevi under the general category of “nevus.”

|  | **Top-1 Accuracy** | **Top-3 Accuracy** | **Top-5 Accuracy** |
| --- | --- | --- | --- |
| **Dermatologist** | 0.56 | 0.70 | - |
| **Medical device** | 0.50 | 0.71 | 0.78 |

### Prospective analysis

The prospective analysis involves evaluating the performance of three different sources: dermatologists assisted by the current medical device Legit.Health, the medical device Legit.Health on its own, and the latest version of the medical device. For the pathological diagnosis assessment, 15 out of the 42 samples without a confirmed pathological examination were excluded due to the fact that these lesions were not excised, though they were included in the diagnosis concordance analisis. The confirmed diagnostic data from this part of the study are presented in Supplementary Table 2.

**Supplementary table 2.** Diseases diagnosed in prospective patients of the study and percentage within the 27 samples analysed.

| **Condition** | **Lesions (n, %)** |
| --- | --- |
| Melanocytic nevus | 12 (44.4) |
| Seborrheic keratosis | 3 (11.1) |
| Basal cell carcinoma | 3 (11.1) |
| Junctional nevus | 3 (11.1) |
| Capillary angioma | 1 (3.7) |
| Dermal melanocytosis | 1 (3.7) |
| Atypical nevus | 1 (3.7) |
| Blue nevus | 1 (3.7) |
| Lentiginous junctional nevus | 1 (3.7) |
| Actinic keratosis | 1 (3.7) |

The malignancy results, as shown in Table 3, demonstrate strong diagnostic performance across all three diagnostic sources, with an AUC exceeding 0.94. In particular, dermatologists achieve an 85% malignancy detection rate when assisted by the Legit.Health medical device. The medical device performs a more conservative diagnosis, leading to the identification of a greater number of malignant pathologies. Sensitivity and specificity statistics were calculated using a threshold of 10, as recommended by the manufacturer. However, as shown in Figure 1, this threshold can be adjusted for the medical device to improve specificity without a loss of sensitivity.

**Table 3.**
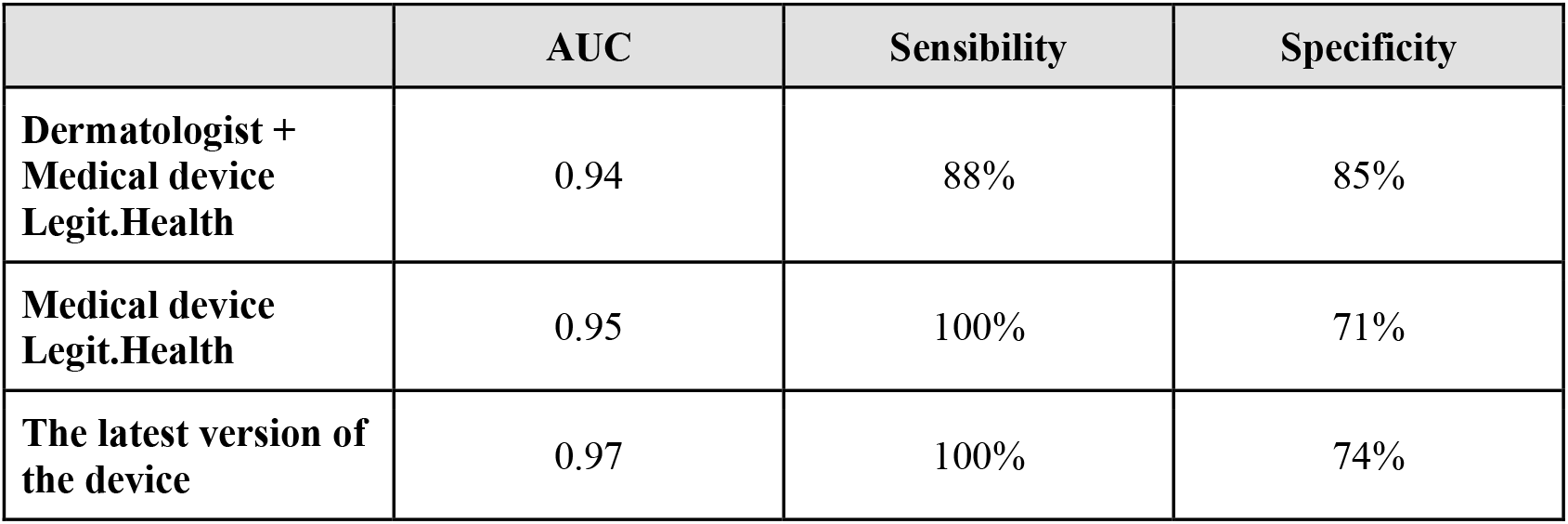
Malignancy diagnostic performance results for prospective patients.

**Figure 1.**
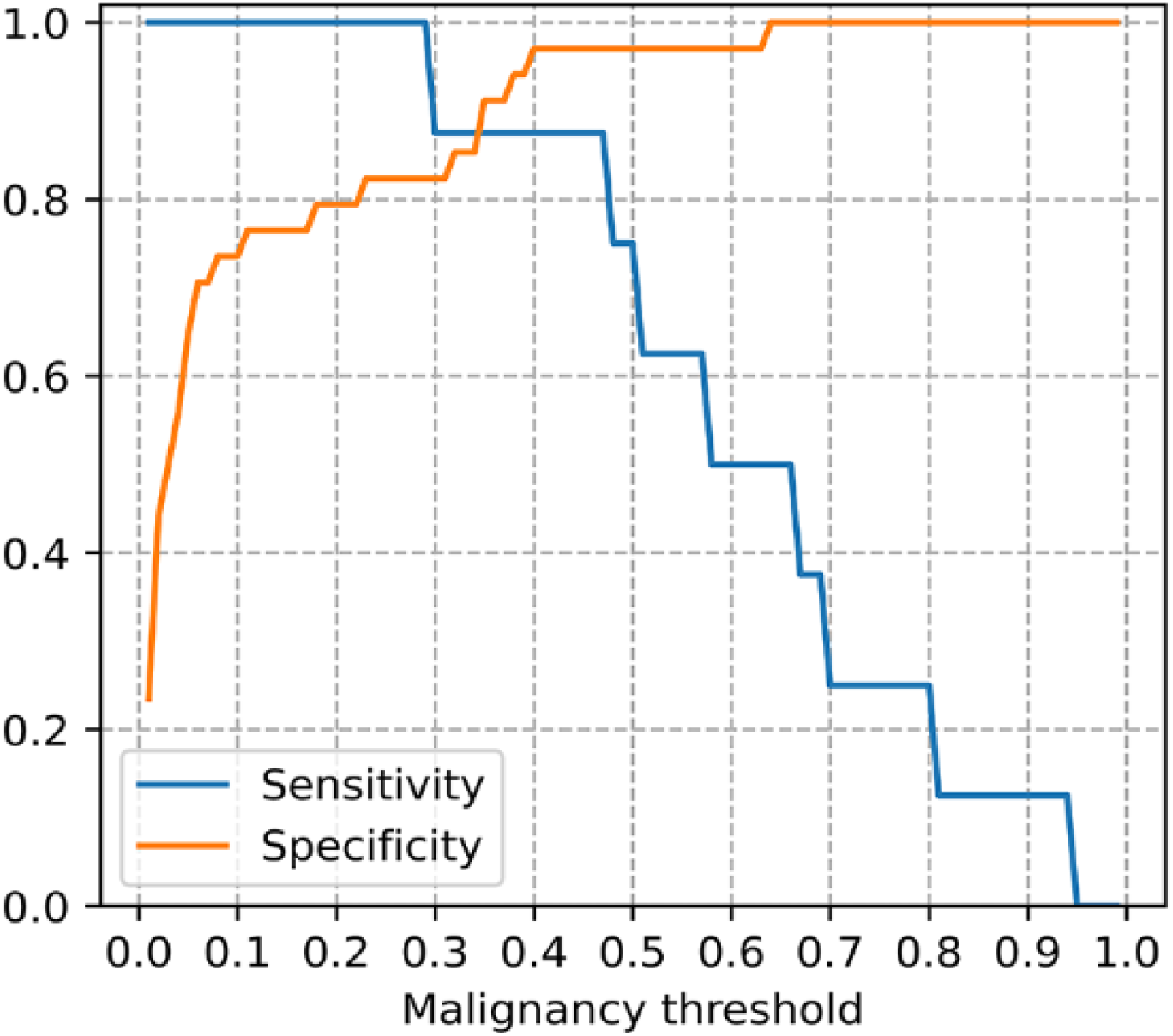
Relation between sensitivity and specificity.

It is important to highlight that the prospective evaluation results include the dermatologist’s recommendation to monitor or excise the lesion. As expected, this recommendation closely aligns with the malignancy index measured by the medical device.

Regarding diagnostic accuracy, we found that the top-K accuracy of dermatologists, when supported by the medical device, is comparable to that of the device operating independently, as shown in Table 4.

**Table 4.** Diagnostic accuracy results for dermatologists and the medical device.

|  | Top-1 Accuracy | Top-3 Accuracy | Top-5 Accuracy |
| --- | --- | --- | --- |
| **Dermatologist +**<br>**Medical device**<br>**Legit.Health** | 0.30 | — | — |
| **Medical device**<br>**Legit.Health** | 0.22 | 0.33 | 0.44 |
| **The latest version of**<br>**the medical device** | 0.26 | 0.37 | 0.52 |

As in the retrospective cases, we found that some samples correspond to various types of naevi. Among these cases, 60–80% of naevi cases are misclassified when attempting to identify the specific histologic subtype of nevus. However, when disregarding the exact histologic subtype, all dermatologists are assisted by the Legit.Health medical device, the device on its own, and the latest version of the medical device makes no diagnostic errors in the naevus samples, leading to improved top-K accuracy. Notably, dermatologists assisted by the medical device nearly triple their accuracy, while the medical device achieves a top-1 accuracy approaching that of dermatologists. The results are presented below in Table 5.

**Table 5.**
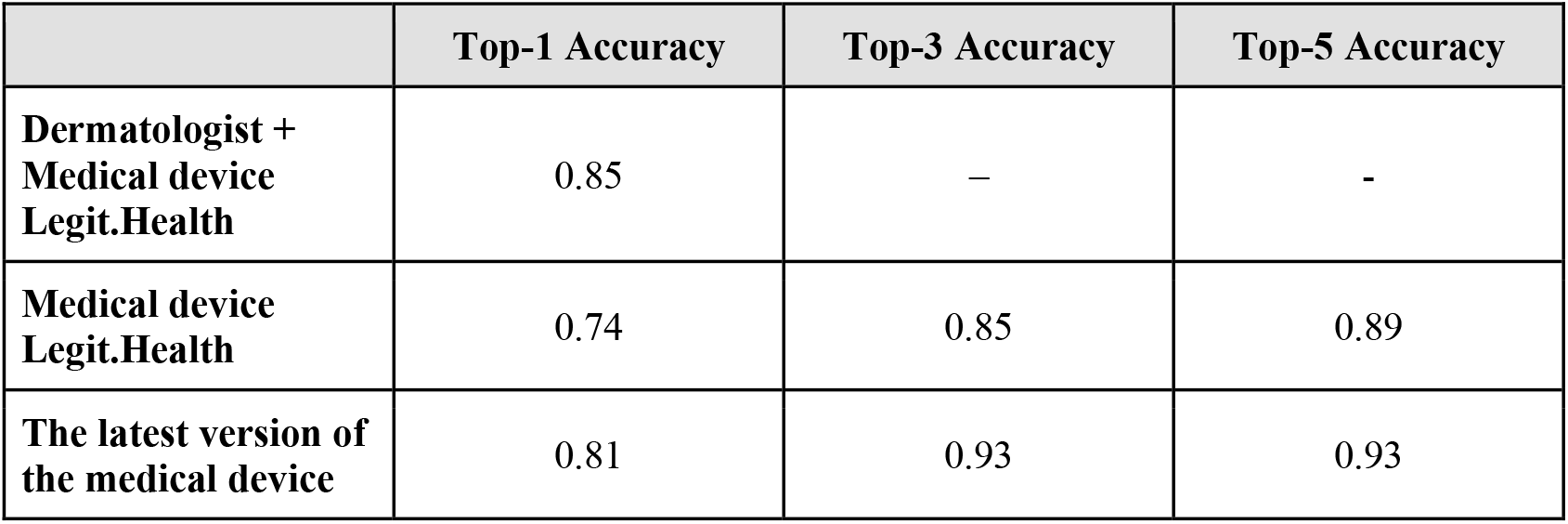
Diagnostic accuracy results for dermatologists and the medical device after grouping different types of naevi under the general category of “nevus.”

The results from the prospective sample demonstrate that the latest version of the current medical device outperforms the Legit.Health medical device across all metrics.

## DISCUSSION

This study demonstrated that the medical device Legit.Health achieved strong performance in detecting malignancy in patients with lesions suspicious of skin malignancy and supporting the diagnosis of cutaneous pathologies using both dermoscopic and clinical images. The results were comparable to those of an expert dermatologist.

The primary objective of the study was to improve the clinical workflow and care process for patients with lesions suspicious of skin malignancy by enhancing diagnosis and severity assessment. In the malignancy analysis of retrospective images, the device achieved an AUC similar to that of dermatologists for malignancy detection. In terms of sensitivity and specificity, sensitivity was nearly identical (86% for dermatologists vs. 81% for the device), while specificity was nearly 20% higher. These malignancy results indicate a greater tendency among dermatologists to diagnose malignant pathologies, which, due to their more conservative approach, could lead to increased resource utilisation in clinical dermatology practice ^14^. It is important to note that similar malignancy detection performance between the medical device and healthcare professionals has been observed in previous studies with other models ^15^, further highlighting its potential to optimise clinical workflow ^16^.

For the prospective image analysis, it was observed that dermatologists benefit from the use of the medical device, demonstrating strong performance in detecting malignant lesions (AUC above 0.94) with high sensitivity and specificity, consistent with the current state of the-art ^17, 18^. Additionally, both dermatologists and the medical device exhibit superior performance when evaluating prospective lesions compared to retrospective ones. This disparity can be attributed to several factors. First, prospective lesions originate from a smaller, more homogeneous dataset consisting mainly of well-characterised pathologies such as seborrhoeic keratosis, basal cell carcinoma, and nevus. In contrast, retrospective lesions show greater variability, including pathologies such as dermatofibroma, melanoma, lentigo, and various carcinomas, which present greater diagnostic challenges. Moreover, the improved performance in the prospective evaluation can be attributed to the fact that dermatologists benefit from the medical device’s assistance, which analyses up to three images, including both, clinical and dermoscopic images, per lesion. By contrast, in the retrospective evaluation, only a single image per lesion (clinical or dermoscopic) was available for analysis.

In this study, a malignancy threshold can be applied to the medical device’s malignancy score to determine when excision may be necessary, providing valuable support for dermatologists’ clinical decision-making. Our analysis revealed that with a malignancy threshold set at 0.40, the medical device can predict the need for excision with 90% accuracy. This information could assist healthcare professionals in deciding patient treatment strategies, such as monitoring, biopsy, or lesion excision, rather than solely providing a specific diagnosis based on clinical assessment. These recommendations are closely related to the malignancy prediction generated by the device ^19^.

Regarding diagnostic support, when the diagnosis of nevus was unified, it was observed in the retrospective image analysis that the device performed similarly to the dermatologist ^10^ and even outperformed them when considering the Top-5 pathologies. However, in the prospective image analysis, dermatologists achieved a diagnostic accuracy of 85% when supported by the medical device, increasing to 93% with the latest version of the device. This result reinforces the device’s intended use as a clinical decision-support tool. It is also important to highlight that Top-1 accuracy was not a primary metric in this study, as the device is specifically designed to always provide the top five predicted classes, aligning with its intended function as a decision support tool. It is also relevant that the differences between different subtypes of melanocytic nevus are mainly histological, thus a clinical-histological correlation does not always exist which should be taken into consideration ^20^.

Additionally, image quality in this study likely influenced the device’s performance. Higher-quality images could have enhanced device functionality and provided more valuable information. Factors such as ensuring that the image is centered on the lesion area and that it is clear and free of hair can improve image quality, thereby preventing any negative impact on the device’s performance. In this way, the medical device is able to filter and discard images of low quality with artifacts that can disturb the quality of the image. Thus, the use of the medical device enables physicians to avoid processing these images.

After analysing the study results across both retrospective and prospective images, it can be concluded that the medical device is a highly valuable tool for improving clinical workflow and patient care in cases of lesions suspicious of skin malignancy. This is achieved through enhanced diagnosis and malignancy prediction, as demonstrated in the prospective image dataset, where dermatologists achieved strong malignancy detection results. This strong performance was achieved despite the inherent bias in the dataset, which only includes lesions deemed sufficiently suspicious to warrant a biopsy. It’s also remarkable the ability of the device to analyse both clinical and dermoscopic images, in the same way as a well-trained physician does. This ability is, in our opinion, a clear advantage of working with a well-trained algorithm for both kinds of images and add significant value to the information available to physicians

In summary, the device represents a cutting-edge solution in dermatological diagnostic support and telemedicine. Its integration of machine learning algorithms, patient-centred approach, and favourable safety profile position it at the forefront of advancements in dermatological technology.

## CONCLUSIONS

The diagnostic capability of the device in distinguishing skin malignancy is on par with that of expert dermatologists, not only in teledermatology but also in in-person consultations, confirming its reliability as a malignancy screening tool within ICD-11 categories. It assists in prioritising patients based on urgency, ensuring appropriate referrals to specialists or relevant clinics, and serves as a diagnostic support tool, thereby facilitating clinical decision-making.

## Data Availability

All data produced in the present study are available upon reasonable request to the authors.

## ACKNOWLEDGEMENTS

We extend our gratitude for the co-funding of this project (DERMATIA: Multicenter Implementation and Validation of an Artificial Intelligence Platform for Clinical Decision Support and Patient Management in Dermatology) by the European Union (NextGenerationEU) through the Public Business Entity Red.es (State Secretariat for Digitalization and Artificial Intelligence, Ministry of Economic Affairs and Digital Transformation) within the framework of the 2021 Call for Grants for research and development projects in artificial intelligence and other digital technologies, and their integration into value chains (C005/21-ED), with file number 2021/C005/00154001.

We thank Clinitax (http://clinitax.eu) for its support and role in securing funding and project management.

